# COVID-19 presenting as anosmia and dysgeusia in New York City emergency departments, March - April, 2020

**DOI:** 10.1101/2020.07.06.20147751

**Authors:** Tina Z. Wang, Jessica Sell, Don Weiss, Ramona Lall

**Affiliations:** Department of Medicine, Division of Infectious Diseases, Weill Cornell Medicine, New York, NY; New York City Department of Health and Mental Hygiene, New York, NY

**Author notes:** **Corresponding Author:** Don Weiss, MD, MPH, Bureau of Communicable Disease, New York City Department of Health and Mental Hygiene, Gotham Center, CN# 22a, 42-09 28th Street, 6th Floor, Queens, New York 11101-4132.

## Abstract

**Background:** Increasing evidence has been emerging of anosmia and dysgeusia as frequently reported symptoms in COVID-19. Improving our understanding of these presenting symptoms may facilitate the prompt recognition of the disease in emergency departments and prevent further transmission.

**Methods:** We examined a cross-sectional cohort using New York City emergency department syndromic surveillance data for March and April 2020. Emergency department visits for anosmia and/or dysgeusia were identified and subsequently matched to the Electronic Clinical Laboratory Reporting System to determine testing results for SARS-CoV-2.

**Results:** Of the 683 patients with anosmia and/or dysgeusia included, SARS-CoV-2 testing was performed for 232 (34%) and 168 (72%) were found to be positive. Median age of all patients presenting with anosmia and/or dysgeusia symptoms was 38, and 54% were female. Anosmia and/or dysgeusia was the sole complaint of 158 (23%) patients, of whom 35 were tested for SARS-CoV-2 and 23 (66%) were positive. While the remaining patients presented with at least one other symptom, nearly half of all patients (n=334, 49%) and more than a third of those who tested positive (n=62, 37%) did not have any of the CDC-established symptoms used for screening of COVID-19 such as fever, cough, shortness of breath, or sore throat.

**Conclusions and Relevance:** Anosmia and/or dysgeusia have been frequent complaints among patients presenting to emergency departments during the COVID-19 pandemic, and, while only a small proportion of patients ultimately underwent testing for SARS-CoV-19, the majority of patients tested have been positive. Anosmia and dysgeusia likely represent underrecognized symptoms of COVID-19 but may have important future implications in disease diagnosis and surveillance.

## Introduction

In March 2020, new anecdotal evidence emerged of anosmia and dysgeusia as important clinical features of COVID-19 infection. A letter by Dr. Claire Hopkins, president of the British Rhinological Society, to ENT UK described patients presenting with anosmia and/or dysgeusia early in the course of COVID-19 infection and occasionally in the absence of respiratory symptoms [1]. These findings have subsequently been reported by researchers in several countries. In a review of neurological manifestations of COVID-19 in China, 12 of 214 (5.6%) patients reported hypogeusia and 11 (5.1%) reported hyposmia [2]. A retrospective survey in Italy found 20 of 59 (33.9%) hospitalized COVID-19 patients reported olfactory or taste dysfunction and 11 (18.6%) reported both [3]. Similar reports have also been published from France, Iran, and South Korea [4,5,6].

These accruing findings have prompted experts and medical societies to propose screening and testing of individuals with anosmia and/or dysgeusia [7]. In response, the American Academy of Otolaryngology-Head and Neck Surgery has collected and analyzed 237 cases of anosmia and/or dysgeusia from around the world. The majority (73%) of patients reported anosmia onset prior to COVID-19 diagnosis and 27% reported no other symptoms [6]. A New York Times article published March 22, 2020 brought further attention to this issue [8]. In late April 2020, the Centers for Disease Control and Prevention (CDC) added “new loss of taste or smell” to the symptoms to watch for in COVID-19 [9].

New York City (NYC) has been the epicenter of the COVID-19 pandemic in the United States. Rapid recognition and triage of cases in emergency departments (EDs) is crucial to the control of the outbreak. In order to establish the role of anosmia and dysgeusia in the identification of COVID-19, we reviewed the prevalence of these chief complaints in ED visits and results of testing for SARS-CoV-2.

### Data and Methods

NYC’s Department of Health and Mental Hygiene (DOHMH) receives ED syndromic surveillance data from all 53 acute care hospital EDs in NYC. By conducting word analysis of chief complaints, we are able to detect new or increasing mentions of individual words. On March 15, 2020, we detected increased mentions of the words “taste” and “smell.”

We analyzed ED data for March and April 2020. Cases were identified by searching chief complaints for mentions of “anosmia”, “ageusia”, “loss of/no/decreased/disturbance/difficulty/unable to taste or smell” utilizing perl regular expressions in R to account for misspellings and differences in sentence structure. Cases were also identified by searching International Classification of Diseases,10th Revision (ICD-10) discharge diagnosis codes for R43 “Disturbances of Smell and Taste.”

To determine whether identified cases underwent SARS-CoV-2 PCR testing and the testing result, we matched cases to the Electronic Clinical Laboratory Reporting System (ECLRS) by using patient name, date of birth, and sex. Definitive name-based matching was performed for 376 (55%) cases. For cases where patient name was unavailable, matching was done using patient date of birth, sex, and ZIP code of residence along with laboratory specimen collection ±7 days from the ED visit date. The latter method was successful in matching cases with an error rate of 8%.

Data compilations, descriptive statistics, and visualizations were conducted using R. The study was evaluated by the NYC DOHMH Institutional Review Board and deemed to be exempt as public health surveillance.

## Results

A total of 715 patients presented to NYC EDs in March and April 2020 with a complaint of anosmia and/or dysgeusia. Of these, 683 from 47 hospitals were NYC residents and were included in our analysis. For comparison, there was an average of 22 visits for anosmia and/or dysgeusia in March and April of the previous 5 years (2015 through 2019). Visits for anosmia and/or dysgeusia increased following the publication of Dr. Hopkin’s letter and the New York Times article **(Figure 1)**, with the majority (72%) of all visits occurring between March 22 and April 11, 2020.

**Figure 1.**
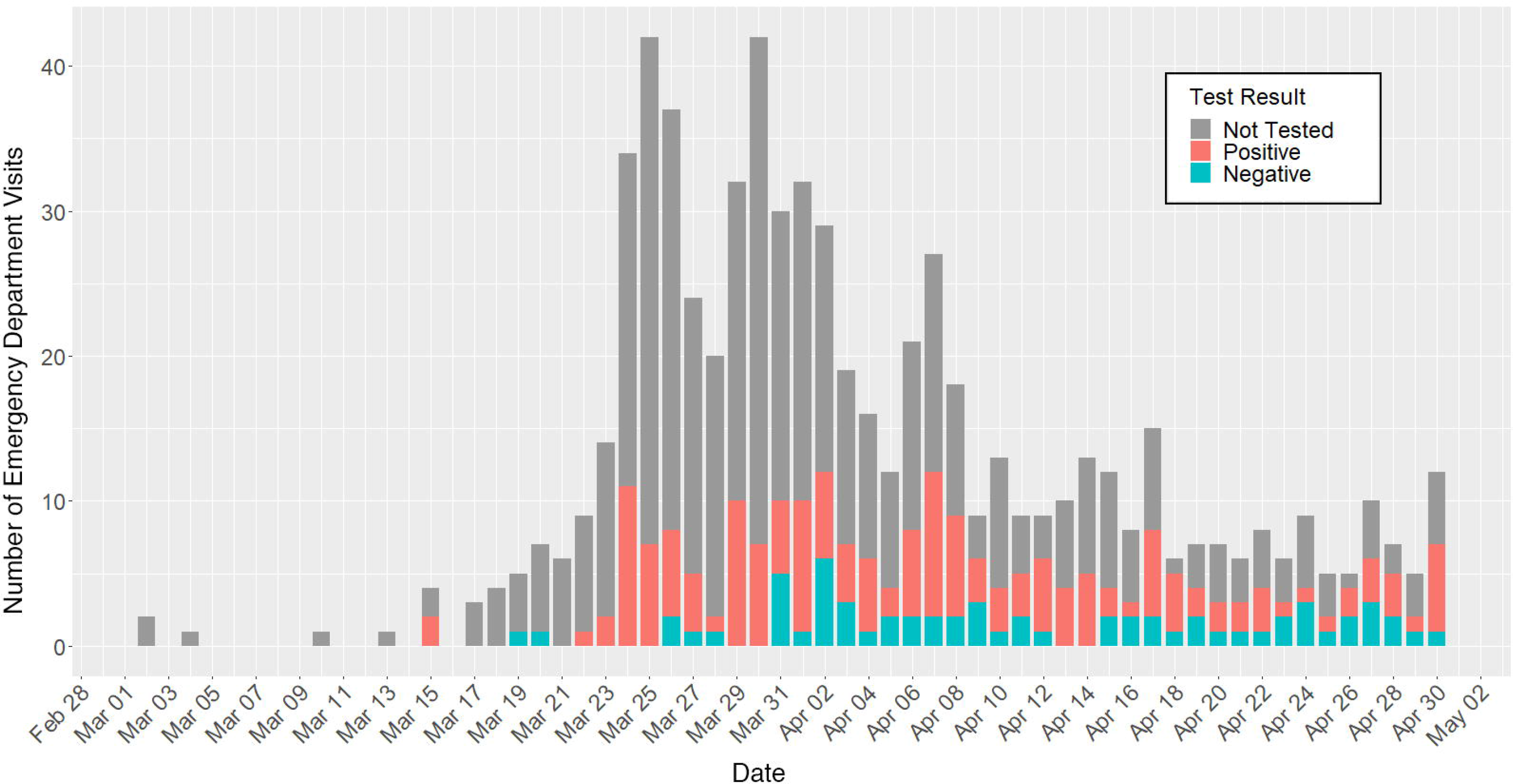
Daily NYC ED Visits for Anosmia and/or Dysgeusia Stratified by Testing.

Of the 683 patients, we identified SARS-CoV-2 results for 232 (34%) and 168 (72%) were confirmed positive. This represents 0.1% of the 175,316 positive SARS-CoV-2 tests in March and April 2020. Demographic and clinical characteristic are summarized in **Table 1**. Median age of all patients was 38 years with the majority under the age of 45 (425, 63%). Female patients comprised a slight majority (366, 54%). Forty-three (6%) of the 683 anosmia and/or dysgeusia were admitted.

**Table 1.** Demographic and Clinical Characteristics.

| **Demographic and Clinical Characteristics** |  |  |  |  |
| --- | --- | --- | --- | --- |
|  | **Total** |  | **COVID-19 positive** |  |
| ***Demographics*** | ***N*** | ***%*** | ***N*** | ***%*** |
| Age | 683 | 38(median) | 168 | 44(median) |
| 3-17 | 18 | 3% | 1 | 1% |
| 18-44 | 407 | 60% | 85 | 51% |
| 45-64 | 206 | 30% | 61 | 36% |
| 65-74 | 43 | 6% | 19 | 11% |
| 75+ | 9 | 1% | 2 | 1% |
| Sex - Female | 366 | 54% | 94 | 56% |
| Borough (County) |  |  |  |  |
| Bronx | 150 | 22% | 28 | 17% |
| Brooklyn | 277 | 41% | 77 | 46% |
| Manhattan | 98 | 14% | 27 | 16% |
| Queens | 123 | 18% | 26 | 15% |
| Staten Island | 35 | 5% | 10 | 6% |
| Race |  |  |  |  |
| Asian | 14 | 2% | 6 | 4% |
| Black or African American | 272 | 40% | 71 | 42% |
| White | 85 | 12% | 23 | 14% |
| Other | 259 | 38% | 56 | 33% |
| Unknown | 53 | 8% | 12 | 7% |
| Ethnicity |  |  |  |  |
| Hispanic | 154 | 23% | 38 | 23% |
| Non-Hispanic | 362 | 53% | 99 | 59% |
| Unknown | 167 | 24% | 10 | 6% |
| **Vital Signs** | **N** | **Median** | **N** | **Median** |
| Temperature | 565 | 98.5 | 147 | 98.6 |
| Temp $\geq 100.4^{\circ}\text{F}$ | 44 | | 19 | |
| Heart Rate | 347 | 87 | 86 | 89.5 |
| Tachycardia ($\geq 100/\text{min}$) | 75 | | 25 | |
| Respiratory rate | 375 | 18 | 90 | 18 |
| Tachypnea ($\geq 20/\text{min}$) | 105 | | 41 | |
| Oxygen Saturation | 487 | 98 | 121 | 98 |
| Hypoxemia ($< 95\%$) | 27 | | 12 | |
| Systolic BP | 401 | 127 | 93 | 126 |
| Diastolic BP | 404 | 80 | 94 | 79 |
| **Symptoms** | **N** | **%** | **N** | **%** |
| No other symptoms | 158 | 23% | 23 | 14% |
| Anosmia alone | 35 | 5% | 8 | 5% |
| Dysgeusia alone | 24 | 4% | 1 | 1% |
| Both anosmia and dysgeusia | 99 | 14% | 14 | 8% |
| Other symptoms |  |  |  |  |
| Cough | 221 | 32% | 68 | 40% |
| Fever | 184 | 27% | 67 | 40% |
| Shortness of Breath | 142 | 21% | 51 | 30% |
| Headache | 85 | 12% | 17 | 10% |
| Nausea, Vomiting, Diarrhea, or<br>Abdominal Pain | 77 | 11% | 23 | 14% |
| Bodyache | 75 | 11% | 23 | 14% |
| Malaise or Fatigue | 73 | 11% | 21 | 13% |
| Chest pain | 67 | 10% | 25 | 15% |
| Sore throat | 44 | 6% | 12 | 7% |
| Nasal congestion | 37 | 5% | 9 | 5% |
| Chills | 35 | 5% | 13 | 8% |
| Dizziness or Syncope | 25 | 4% | 7 | 4% |

One hundred and fifty-eight (23%) patients presented with anosmia or dysgeusia in the absence of any other reported symptoms. Of these, 35 were tested for SARS-CoV-2 and 23 (66%) tested positive. Anosmia alone was seen in 35 (5%) of COVID-19 visits and 8 (5%) positive cases, while dysgeusia alone was the presenting complaint in 24 (4%) COVID-19 visits and 1 (1%) positive case. The remaining patients reported anosmia and/or dysgeusia in addition to at least one other symptom. Only 349 (51%) of all patients and 106 (63%) of those who tested positive presented with one or more CDC-established symptoms recommended for screening of COVID-19 disease (cough, fever, shortness of breath, or sore throat) **(Figure 2)**.

**Figure 2.**
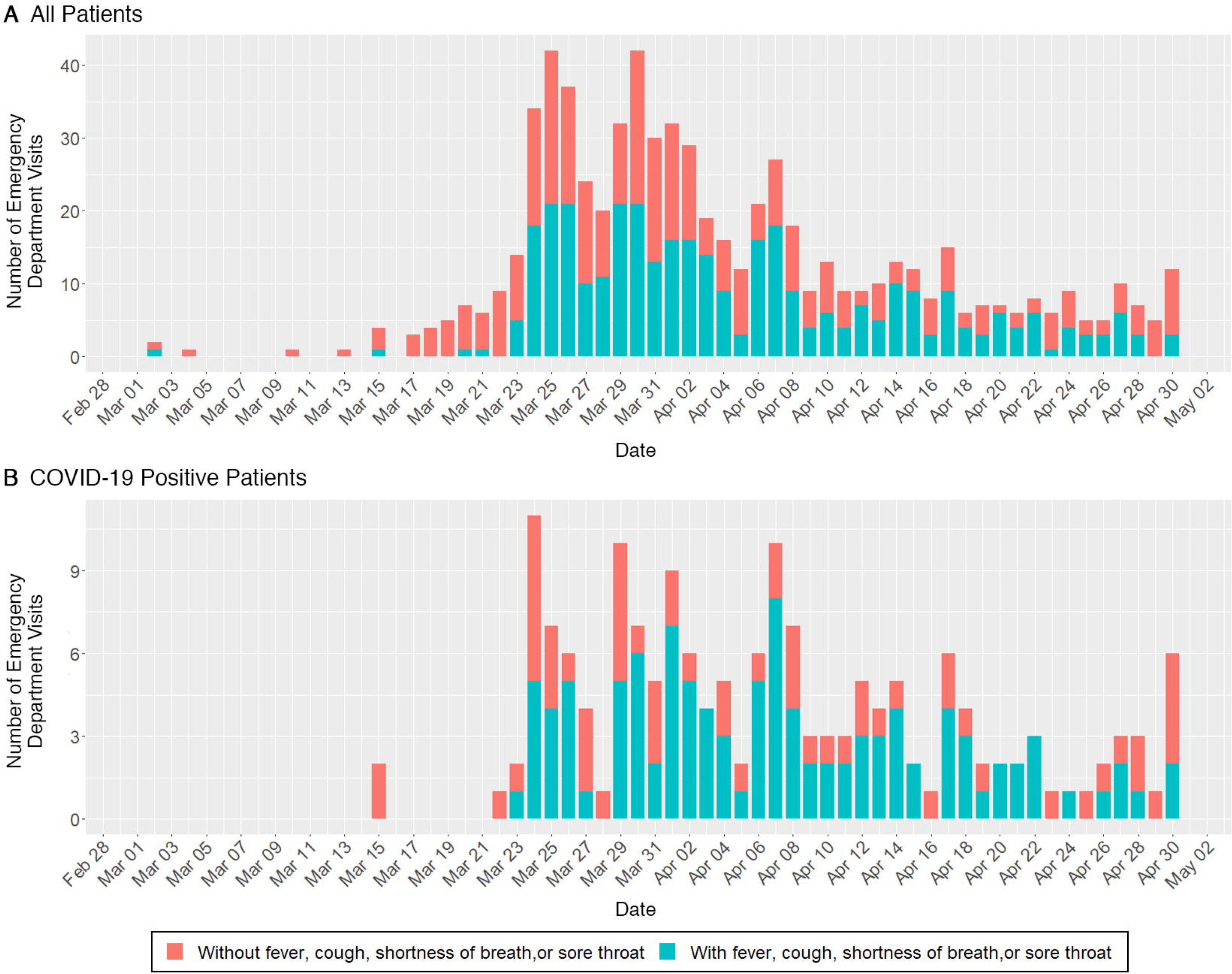
Daily NYC ED Visits for Anosmia and/or Dysgeusia Stratified by Presence of Other Symptoms.

Among the 238 patients who underwent SARS-CoV-2 testing, 16 (7%) had testing performed prior to presenting with anosmia and/or dysgeusia symptoms and 124 (52%) received testing during the same ED visit. The remaining 98 (41%) underwent testing at a subsequent healthcare encounter a median of 7 days after their initial anosmia and/or dysgeusia ED visit and 56 (57%) were found to be SARS-CoV-2 positive.

## Discussion

Anosmia and dysgeusia have been uncommon complaints of persons presenting to NYC EDs in previous years. Our data show that reports of these symptoms dramatically increased in late March 2020 yet testing of patients with these complaints remained low. While only 34% of these patients underwent testing for COVID-19, the majority (72%) of those tested were positive for the infection. Of those tested, only 52% appeared to be tested as part of their ED visit. It is possible that patients presenting to EDs with anosmia and dysgeusia were milder cases who did not meet stricter testing criteria. Our study demonstrates that among those that did get tested there was a high percentage positive, suggesting anosmia and dysgeusia are reliable symptoms for COVID-19, and confirming findings of prior studies. Awareness of these symptoms as potential signs of COVID-19 only recently emerged with the modification of CDC’s symptom list in late April. With further knowledge and expanded future testing, the proportion of persons with anosmia and/or dysgeusia may represent an even larger population than our study was able to capture.

Our findings support previous reports that anosmia and/or dysgeusia may be a frequent complaint among patients with COVID-19 and may present in the absence of other symptoms [1, 3, 6]. A significant proportion presented without classic respiratory illness symptoms of cough, fever, and shortness of breath, and were not tested for SARS-CoV-2. As our study shows, this presentation may be especially important in younger, female patients, which is in contrast to characteristics of COVID-19-infected patients in the general population who are older and predominantly male [10].

Some experts have projected that COVID-19 may eventually assume a seasonal pattern of outbreaks similar to influenza. If a future resurgence of COVID-19 occurs during influenza season, discrimination of the two infections may be hindered by the commonality between their presenting symptoms. Anosmia and dysgeusia, which some propose to be caused by direct damage to olfactory and gustatory receptors by SARS-CoV-2, may be key in distinguishing COVID-19 from other influenza-like illnesses in ED settings [6,11]. Furthermore, the tracking of anosmia and dysgeusia complaints may play an important role in alerting public health agencies to impending outbreaks.

Our study was limited by its nature as a surveillance study. The data collected on symptoms were drawn from ED medical records rather than patient interview and may not reflect all of a patient’s complaints. Six hospitals did not contribute any patients. This could either represent a lack of COVID-19 suspect cases at these hospitals or failure of the syndrome definition to capture them. Additionally, patient name for matching to laboratory records was only available for 55% of our cases. The remaining cases utilized other demographic data to match to SARS-CoV-2 test results in ECLRS, which likely led to a small number of mismatches. Lastly, shortages in testing availability and strict testing criteria throughout NYC in March and April of 2020 likely limited the ability to determine the true number of SARS-CoV-2 positive cases.

To date, understanding of the pathogenesis, prevalence, and importance of these symptoms remain incomplete. As our knowledge of COVID-19 evolves, we hope that findings from this study will improve our understanding of the significance of non-respiratory symptoms in COVID-19 and help to inform public health decisions.

## Data Availability

The data that support the findings of this study are available from the corresponding author, upon reasonable request.

